# Comprehensive evaluation of AT(N) imaging biomarkers for predicting cognition

**DOI:** 10.1101/2024.11.25.24317943

**Authors:** Tom Earnest, Braden Yang, Deydeep Kothapalli, Aristeidis Sotiras, the Alzheimer’s Disease Neuroimaging Initiative

## Abstract

**Background and Objectives:** Imaging biomarkers enable *in vivo* quantification of amyloid, tau, and neurogenerative pathologies that develop in Alzheimer’s Disease (AD). Interest in imaging biomarkers has led to a wide variety of biomarker definitions, some of which potentially offer less predictive value than others. We aimed to assess how different operationalizations of AD imaging biomarkers affect prediction of cognition.

**Methods:** We included individuals from ADNI who underwent amyloid-PET ([^18^F]-Florbetapir), tau-PET ([^18^F]-Flortaucipir), and volumetric MRI imaging. We compiled a large collection of imaging biomarker definitions (42 in total) spanning different pathologies (amyloid, tau, neurodegeneration) and variable types (continuous, binary, non-binary categorical). Using cross-validation, we trained regression models to predict neuropsychological performance, both globally and across different subdomains (Phenotype Harmonization Consortium composites), using different combinations of biomarkers. We also compared these biomarker models to support vector machines (SVMs) trained to predict cognition directly from imaging regions of interest. In a subsample of individuals with CSF biomarker readouts, we repeated experiments comparing the accuracy of models using imaging and fluid biomarkers. Additional analyses tested the predictive strength of imaging biomarkers when limited to specific clinical stages of disease (cognitive unimpaired vs. impaired) and when modeling longitudinal cognitive change.

**Results:** Our sample included 490 people (247 female) with a mix of no impairment (n=288), mild impairment (n=163), and dementia (n=39). While almost all biomarkers tested were predictive of cognitive performance, we observed substantial variability in accuracy, even for measures of the same pathology. Tau biomarkers were the single most accurate single predictors, though combination of biomarkers spanning multiple pathologies were more accurate overall. SVM models were generally more accurate than models using traditional biomarkers. Incorporating continuous or non-binary categorical biomarkers was beneficial only for tau and neurodegeneration, but not amyloid. Patterns of results were largely consistent when considering different clinical stages of disease, neuropsychological domains, and longitudinal cognition. In the CSF subsample (n=246), imaging biomarkers strongly outperformed CSF versions for cognitive prediction.

**Discussion:** We demonstrated that different imaging biomarker definitions can lead to variability in downstream predictive tasks. Researchers should consider how their biomarker operationalizations may help or hinder the assessment of disease severity.

## Introduction

The modern biological definition of Alzheimer’s Disease (AD) relies on biomarkers.^1–3^ Biomarkers can accurately quantify pathobiological disease processes which are specific to AD, particularly the aggregation of amyloid-beta (Aβ) plaques and the neocortical spread of tau neurofibrillary tangles. Importantly, biomarkers can detect and measure these pathologies prior to symptomatic onset. Because of their capabilities, biomarkers have been used in a variety of research settings including disease classification^4^, cognitive forecasting^5^, subtype identification^6^, clinical trial stratification^7^, disease staging^8,9^, and more. Moreover, biomarkers are becoming increasingly important for clinical management of AD^2^. For instance, recently approved anti-Aβ treatments for AD require the presence of Aβ-pathology as assessed by biomarkers.

Interest in biological AD assessment has led to the creation of many AD-sensitive biomarkers which vary in terms of modality, underlying pathology, and statistical formulation. Idiosyncrasies of biomarker definitions may result in unwanted variability when applied for clinical and research uses. For example, estimated cut points for PET and CSF biomarker dichotomization are fairly application specific ^10–12^, and different approaches to pathological thresholding result in considerable variability for group assignment^13–16^. Less is known, however, about how variability in biomarker definitions affects prediction of cognition in AD. Identifying which specific biomarkers are most predictive of cognitive trajectories, particularly at different stages of disease, can provide insight into biological mechanisms of AD. Moreover, precise cognitive decline predictions are valuable for identifying candidates for early therapeutic interventions and for establishing meaningful cognitive endpoints in clinical trials. Despite these implications, investigations into the ramifications of different biomarker operationalizations remain limited. One previous study found that different biomarker definitions varied in their ability to predict longitudinal Mini-Mental State Examination (MMSE) scores, and that dichotomization hindered predictive power of some biomarkers relative to continuous values.^13^ Similar analyses in separate cohorts with additional cognitive measures are needed to confirm and extend these findings, particularly to establish optimal biomarker combinations for both prognostic accuracy and mechanistic insight.

Here, we developed a comprehensive set of neuroimaging measures (42 in total) covering the AD core biomarkers Aβ (A) and tau (T), as well as non-specific biomarkers of neurodegeneration ((N)). We systematically evaluated how different categories of biomarkers and individual variants differ in their ability to predict different cognitive outcomes. While we focused on cross-sectional cognition, we also extended analyses to measures of prospective longitudinal cognition and neuropsychological domains. We additionally incorporated machine learning to test how traditional biomarker approaches compare to methods which can detect more complex, multivariate patterns in imaging data. Finally, we tested multiple CSF biomarkers (20 definitions spanning 4 analytes) and compared their performance with imaging alternatives.

## Methods

### Participants

We selected a baseline, cross-sectional sample of Alzheimer’s Disease Neuroimaging Initiative (ADNI) participants with tau-PET, Aβ-PET, and structural MRI imaging data. Exclusion criteria were gaps between scans of greater than 1 year or missing values for any of the following variables: age, sex, *APOE* genotype, Clinical Dementia Rating® (CDR) status^17^, Phenotype Harmonization Consortium (PHC) cognitive composite scores^18^.

### Standard protocol approvals, registrations, and patient consents

All participants provided informed written consent for participating in ADNI. Study protocols were approved by site-specific institutional review and ethical boards.

### Image acquisition and processing

Detailed descriptions of imaging protocols are provided on the ADNI website^19^. Briefly, T1-weighted MRI acquisitions were collected on 3T scanners using an accelerated MPRAGE sequence. Aβ-PET scans were acquired 50-70 minutes (4 frames × 5 minutes) after a 370 MBq (± 10%) injection of [^18^F]-Florbetapir. Tau-PET scans were acquired 75-105 minutes (6 frames × 5 minutes) after a 370 MBq (± 10%) injection of [^18^F]-Flortaucipir.

We accessed processed MRI and PET derivatives generated by the ADNI PET Core. A Freesurfer (v7.1.1) processing pipeline was applied to MRI scans to generate gray matter volumes within regions of interest (ROIs) of standard subcortical^20^ and cortical atlases^21^. PET standardized uptake value ratios (SUVRs) were generated for these same ROIs after coregistration of each PET image to a contemporaneous MRI scan. Our analyses incorporated unilateral values from 68 cortical and 14 subcortical gray matter regions. Volumes were standardized relative to the intracranial volume. Aβ-PET uptakes were standardized to a whole cerebellum ROI, while tau-PET uptakes were standardized relative to an ROI containing inferior cerebellar gray matter^22^.

Partial volume corrected (PVC) PET uptakes were available for tau (Geometric Transfer Matrix approach^23,24^) but not for Aβ. We used uncorrected SUVR values for most experiments, but we repeated some experiments with PVC-corrected tau SUVRs to evaluate the effect of PVC on cognitive prediction accuracy.

### Cognitive and clinical assessments

Cognition was assessed using composite scores developed by the PHC^18^. We averaged the memory (PHC_Memory_), executive functioning (PHC_EF_), visuospatial (PHC_Visual_), and language (PHC_Language_) composites to create one global cognitive composite (PHC_Global_). Composites are unitless factor loadings, with lower scores corresponding to more impairment.

CDR was used as a measure of dementia severity^17^. Subjects were assigned to the following groups based on CDR status: cognitively unimpaired (CU, CDR=0) or cognitively impaired (CI, CDR>=0.5).

### Image-based biomarker definitions

We implemented a variety of biomarker definitions to use for predicting cognition. A full list of the biomarker definitions tested is provided in eTable 1. Biomarkers were categorized based on pathology (AT(N)) and variable type (binary [BIN], non-binary categorical [CAT], continuous [CON]). Lists of atlas regions used to form composites are provided in eTable2.

Continuous variables consisted of scalar MRI (volume) or PET (SUVR) measures in standard composite ROIs. For Aβ, continuous measures of Aβ included the average SUVR in a cortical summary region^25,26^ (Aβ composite) and Centiloid^27^. Centiloids were provided by ADNI and derived from the Aβ composite using previously validated equations^28^. Continuous tau measures included the average uptakes in a meta-temporal (MT) composite region^11^ and uptakes in ROIs corresponding to progressive Braak stages^29,30^ (Braak I, Braak III/IV, Braak V/VI). Braak II was omitted due to off-target binding issues with flortaucipir^31,32^. Hippocampal volume and volume of the MT region were included as continuous assessments of neurodegeneration.

Binary predictors consisted of dichotomized versions of the continuous predictors listed above. There were three main methods tested for binarizing continuous variables: previously published cutoffs, Z-scoring, and Gaussian mixture modeling (GMM). Previously published cutoffs were included for the Aβ composite at the following SUVRs: 1.11^33^, 1.24^34^, 1.30^11^, 1.42^11^. We also tested Centiloid cutoffs (15, 20, 25, 30) based on ranges reported in previous literature^28,35,36^. Z-scoring and GMMs were included as data-driven approaches for deriving cutoffs. These methods were applied to the Aβ composite SUVR, MT tau SUVR, MT volume, and hippocampal volume. Z-scores for each variable were computed relative to CU, Aβ-negative individuals (using an SUVR cutoff of 1.11 applied to the Aβ composite to determine Aβ-negativity, as recommended by the ADNI PET Core). Z-scores were dichotomized using cutoffs of 2 and 2.5 standard deviations away from the CU, Aβ-negative mean value. GMM binarization was implemented by fitting two-component Gaussian mixtures to the distribution of continuous variables. A cutoff point was estimated as the curve intersection between the fitted Gaussians. GMMs were omitted for hippocampal volume, due to a lack of bimodal distribution.

Non-binary categorical biomarkers consisted of quartiles, binarization with an indeterminate zone, and staging systems. Quartiles were computed by binning continuous values at the 25^th^, 50^th^, and 75^th^ percentiles. Binarization with an intermediate zone (BIZ) was used to model the uncertainty of assigning individuals who display biomarker values near the cutoff threshold^2^. BIZ was implemented with a GMM, where individuals were marked as uncertain if they showed less than 60% probability of being assigned to either Gaussian component.

Staging systems were included as non-binary categorical measures which assign disease severity grades based on the spatial extent of Aβ or tau pathology. For Aβ, we applied two previously published staging models^9,37^. For tau, we implemented two versions of Braak staging with different granularities: Braak staging (6) (I, III, IV, V, VI) and Braak staging (3) (I, III/IV, V/VI). Detailed description of the staging procedures for each of these systems is provided in the eMethods.

### CSF-based biomarkers

We identified a subsample of individuals who had CSF immunoassays within 1 year of imaging. CSF samples were analyzed with Roche Elecsys kits sensitive to Aβ_42_, Aβ_40_, total-tau (tTau), and tau phosphorylated at threonine 181 (pTau181). CSF processing was administered by the ADNI Biomarker Core at the University of Pennsylvania.

CSF concentrations of analytes were grouped into biomarker categories as previously recommended^14,151^ (Aβ: Aβ_42_, Aβ_40_, Aβ_42_/Aβ_40_, tau: pTau181, neurodegeneration: tTau). Raw concentrations were included as continuous measures. GMMs were used to define binary versions of CSF biomarkers. BIZ and quartiles were used to generate non-binary categorical versions.

### Statistical analyses

We ran a series of cross-validated modeling experiments to assess how different biomarker definitions compared in their ability to model cognition. Complete details of these experiments are provided in the eMethods. Briefly, we used linear regression models to predict PHC_Global_ using single or multiple biomarker definitions. All regression models also included covariates of age, sex, and *APOE* E4 carriership. Models which only included these covariates were included as controls. Models with single biomarkers were used to test (1) if all included biomarker definitions improved cognitive prediction accuracy and (2) which individual definitions were most predictive of cognitive impairment. Next, we developed linear models combining multiple biomarker definitions as predictors of PHC_Global_. To limit comparisons for these models, biomarkers were grouped based on the underlying pathology (AT(N)) and the variable type (binary, non-binary categorical, continuous), with nested cross-validation used to select the best predicting definition within each group. These models were used to test (3) if combination of biomarkers improved prediction accuracy, and (4) if models incorporating continuous or non-categorical binary biomarkers outperformed models with binary biomarkers. Next, we trained support vector machines (SVM) to predict PHC_Global_ from regional biomarker values to test if (5) multivariate modeling of AD pathology could improve prediction accuracy beyond that of pre-defined biomarker definitions. SVMs were trained with both linear and non-linear kernels with a grid search to select optimal hyperparameters (see eMethods).

All cross-validation experiments had 10 outer folds and were repeated 10 times to generate 100 out of sample error estimates for each tested model. Model error was assessed using root mean squared error (RMSE). Boxplots included in the Results show distributions of the 100 out-of-sample RMSE measurements from trained models. To statistically evaluate differences in accuracy for comparisons of interest, we used Nadeau-Bengio t-tests. The Nadeau-Bengio t-test includes a bias correction for the interdependency of out-of-sample error estimates when using repeated, cross-validated designs^38,39^. All tests were corrected for multiple comparisons with a false discovery rate method^40^. We investigated feature importance by plotting the distribution of selected biomarkers across folds, the magnitude of the coefficients on biomarkers in linear regression models, and the covariance corrected weights for SVM models^41^ (see eMethods). Additionally, we visualized the distribution of model-selected SUVR cutoffs for binary Aβ-and tau-PET biomarkers.

Finally, we ran a series of additional cross-validated experiments using alternative features, target variables, or clinical disease states. Specific experiments were as follows: (a) predicting the prospective slope of PHC_Global_ instead of the cross-sectional value, (b) predicting neuropsychological domains instead of PHC_Global_, (c) using PVC tau data instead of non-PVC, (d) using CSF-based biomarkers instead of imaging-based versions, and (e) using only CU or CI individuals for model selection and out-of-sample evaluation. For (a), we only included individuals who had longitudinal cognitive measurements following baseline. To estimate longitudinal change in PHC_Global_, linear mixed effect models were fit to model longitudinal scores following the baseline assessment. Models were fit with random slopes and intercepts for participants.

## Data availability

Data used in the preparation of this article were obtained from the Alzheimer’s Disease Neuroimaging Initiative (ADNI) database (adni.loni.usc.edu). The ADNI was launched in 2003 as a public-private partnership, led by Principal Investigator Michael W. Weiner, MD. The primary goal of ADNI has been to test whether serial MRI, PET, other biological markers, and clinical and neuropsychological assessment can be combined to measure the progression of MCI and early AD. For up-to-date information, see www.adni-info.org. All data used in this study are accessible from ADNI following formal data usage agreements. Data were downloaded on May 10^th^, 2024. All R (v4.4.0) and Python (v3.10) code for this project will be shared at the following repository: https://github.com/sotiraslab/earnest_ad_biomarker_modeling.

## Results

### Sample characteristics

We selected 490 individuals with baseline biomarker imaging (Table 1). The cohort consisted of a mix of individuals with no cognitive impairment (CDR=0, n=288), very mild dementia (CDR=0.5, n=163), and mild to severe dementia (CDR>0.5, n=39). We observed significant differences in the distribution of age (p=0.001), sex (p=0.005), Aβ-burden (Centilioid, p<0.001), and PHC_Global_ (p<0.001) across dementia status. Mean age and Aβ-burden increased with dementia status, while PHC_Global_ decreased. Relatively more females were observed in the CU group (56.6%) than those with very mild (41.1%) or mild to severe dementia (43.6%). *APOE* E4 status was not significantly different across groups (p=0.200).

**Table 1:** Sample characteristics. The last column shows p-values for significance tests comparing distributions of variables across dementia status groups (CDR). Chi-squared tests of association were used for categorical variables (sex, *APOE* status) and one-way ANOVAs were used for continuous variables (age, Centiloid, PHC_Global_).

|  | CDR=0.0 | CDR=0.5 | CDR=1.0+ | p-value |
| --- | --- | --- | --- | --- |
| **n** | 288 | 163 | 39 |  |
| **Age** | 73.84 (7.44) | 75.78 (8.46) | 78.29 (8.58) | **0.001** |
| **Sex (M/F)** | 125/163 | 96/67 | 22/17 | **0.005** |
| **APOE E4+** | 99 (34.4%) | 62 (38.0%) | 19 (48.7%) | 0.200 |
| **Centiloid** | 19.90 (36.10) | 45.57 (54.95) | 78.41 (49.61) | **<0.001** |
| **PHC<sub>Global</sub>** | 0.81 (0.36) | 0.32 (0.46) | -0.42 (0.54) | **<0.001** |

We also selected subsamples of individuals who had longitudinal PHC_Global_ assessments following baseline (n=383) and those who had CSF biomarker measurements as well as imaging (n=246). Characteristics of these samples are shown in eTables 4 and 5, respectively.

### Assessment of modeling performance for biomarkers

Relative to a control model which included covariates (RMSE=0.531), almost all tested biomarkers led to a significant improvement in prediction accuracy for modeling cognitive scores (Figure 1). The only exception was hippocampal volume binarized at -2.5 Z-scores (RMSE=0.525 [0.06], p=0.09). While these results indicated that most biomarker definitions provided some predictive value, gains in performance were not equal across pathologies and variable types (range in RMSE reduction: 3.4-21.1%). Tau biomarkers led to the largest improvements in accuracy, with 9/10 of the best performing biomarkers being tau-based. Furthermore, SVM models which were trained on regional pathology were more accurate than linear models using single biomarker definitions. The tau SVM was the best performing model overall (RMSE=0.419 [0.05], p<0.001), while the Aβ SVM (RMSE=0.472 [0.06], p<0.001) and volume SVM (RMSE=0.452 [0.05], p<0.001) were the best performing Aβ and neurodegeneration models, respectively. Outside of SVMs, the best performing models for each AT(N) category were the Aβ SUVR binarized at 1.24 (RMSE=0.492 [0.06], p<0.001), continuous MT tau SUVR (RMSE=0.440 [0.05], p<0.001), and continuous MT volume (RMSE=0.471 [0.05], p<0.001).

**Figure 1:**
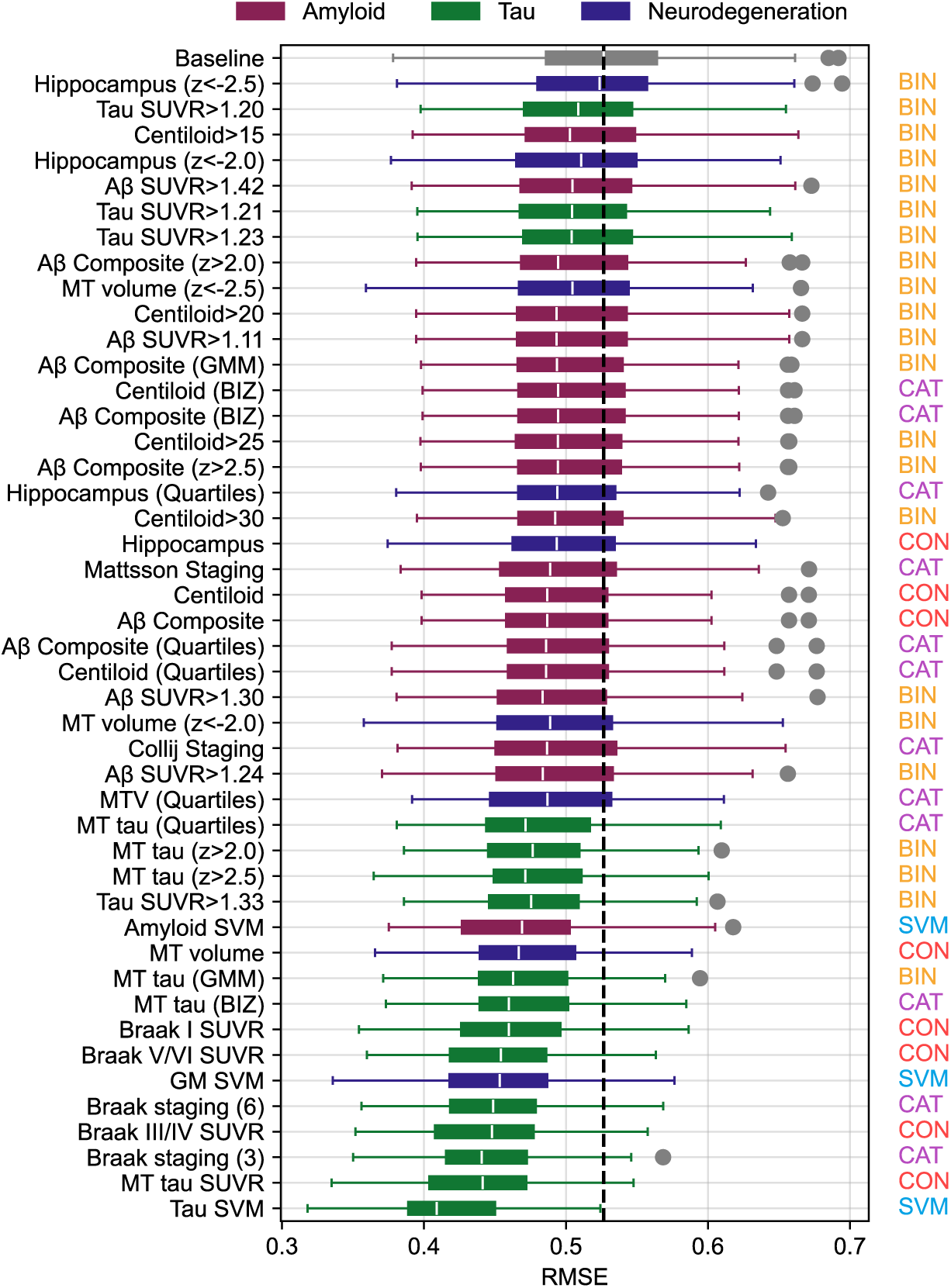
Boxplots showing performance of individual biomarkers for predicting PHC_Global._ Values plotted are RMSEs taken from out of sample predictions for 100 cross-validation instances (lower value is more accurate). The baseline model only included covariates as independent variables (mean performance for this model indicated by the dotted line). All other models included the same covariates and a single amyloid (maroon), tau (green), or neurodegeneration (blue) biomarker. Labels on the right indicate the variable type of each biomarker (BIN=binary, CAT=non-binary categorical, CON=continuous, SVM=support vector machine). All models exhibited significantly lower RMSE than the baseline model, except for hippocampal volume binarized at -2.5 Z-scores [Hippocampus (z<-2.5)].

### Combination of biomarkers

Our next experiments applied a model selection to identify the best performing biomarker predictors based on AT(N) category and variable type. We observed that all biomarker varieties caused a reduction in error over the covariate-only model (Figure 2A; mean RMSEs: Covariates=0.531, A_BIN_=0.498, A_CAT_=0.495, A_CON_=0.495, T_BIN_=0.470, T_CAT_=0.443, T_CON_=0.440, N_BIN_=0.495, N_CAT_=0.492, N_CON_=0.471; all p<0.01). Like our experiments (Figure 1), benefits were largest for tau predictors relative to Aβ and neurodegeneration.

**Figure 2:**
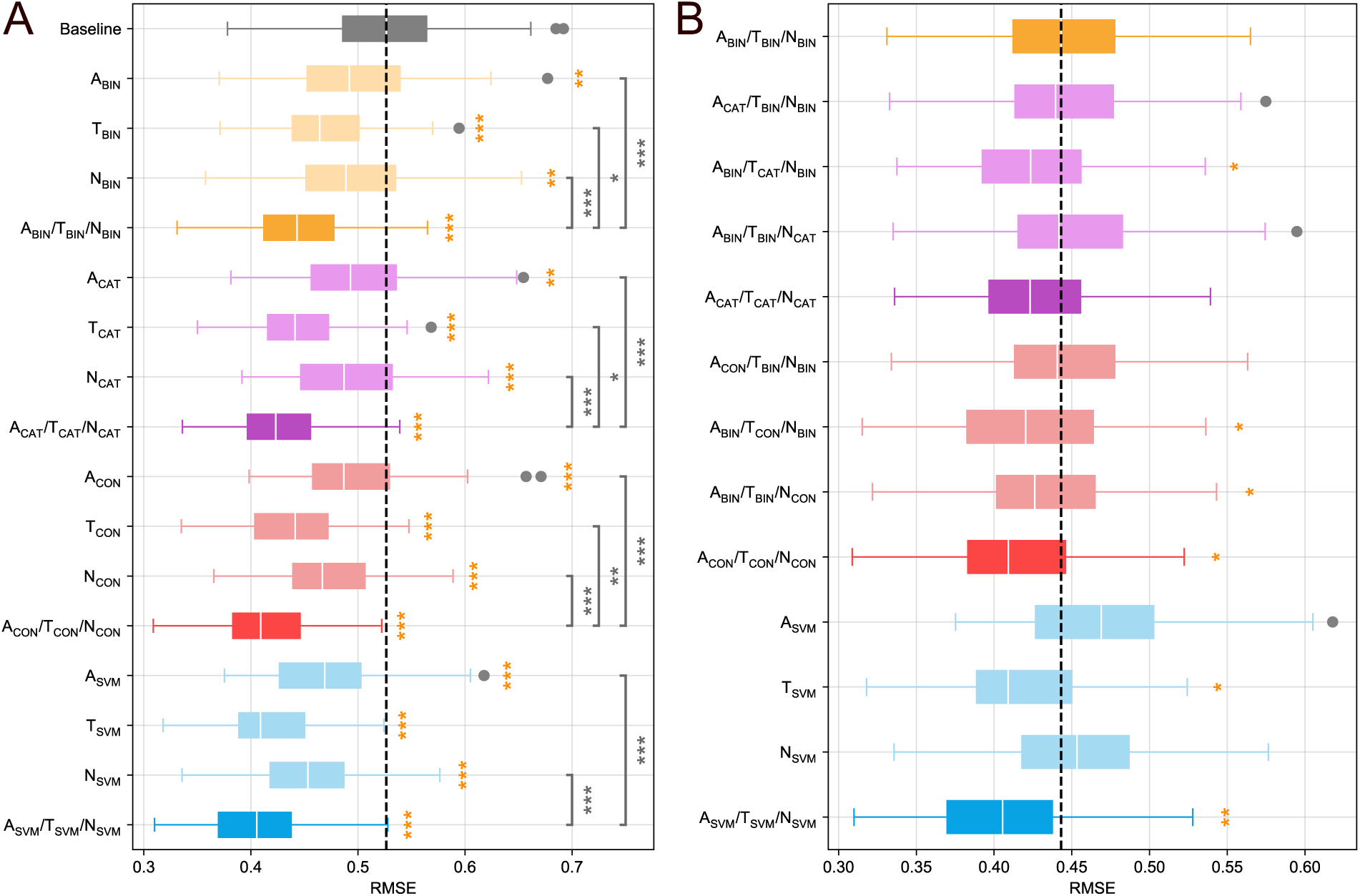
Boxplots showing RMSE performance of combination biomarkers for predicting global neuropsychological performance (PHC_Global_). **A.** Individual and combination biomarker models are compared against a baseline model using only covariates (mean performance indicated by dotted line) to predict PHC_Global._ **B.** Combination biomarker models with non-binary variable types are compared against a baseline model with binary biomarker definitions (mean performance indicated by dotted line). In both panels, colors are used to indicate the variable type of included biomarkers (yellow: binary, purple: non-binary categorical, red: continuous, blue: SVM). Lighter coloring indicates models which only have a single pathology assessment, while darker coloring indicates models which have Aβ, tau, and neurodegeneration biomarkers. Gold stars indicate a significant improvement in accuracy relative to the topmost model. Gray stars and bars highlight significant pairwise differences between individual models. Statistical results are derived from Nadeau-Bengio t-tests with correction for multiple comparisons (*p<0.05, **p<0.01, ***p<0.001).

Combination models which included assessments for all AT(N) categories generally outperformed models with only one category included. All combination models were more accurate than the covariate-only model in predicting global cognition (mean RMSEs: A_BIN_/T_BIN_/N_BIN_=0.446, A_CAT_/T_CAT_/N_CAT_=0.428, A_CON_/T_CON_/N_CON_=0.415, A_SVM_/T_SVM_/N_SVM_=0.405, all p<0.001). Additionally, models which combined biomarkers resulted in significantly higher accuracy than models which only included one pathology assessment (Figure 2A). The benefit of combination was evident for binary (A_BIN_/T_BIN_/N_BIN_ vs. A_BIN_: t=3.96, p<0.001; vs. T_BIN_: t=2.61, p<0.05; vs. N_BIN_: t=3.92, p<0.001), non-binary categorical (A_CAT_/T_CAT_/N_CAT_ vs. A_CAT_: t=4.46, p<0.001; vs. T_CAT_: t=2.22, p<0.05; vs. N_CAT_: t=4.31 p<0.001), and continuous (A_CON_/T_CON_/N_CON_ vs. A_CON_: t=5.22, p<0.001; vs. T_CON_: t=3.34, p<0.01; vs. N_CON_: t=4.48 p<0.001) biomarkers. The combination SVM outperformed the Aβ (t=5.29, p<0.001) and gray matter (t=3.62, p<0.001) SVMs, but not the tau SVM (t=1.56, p=0.12), indicating that the improved accuracy of the multimodal SVM may be primarily driven by tau.

Direct comparison of biomarkers based on variable types indicated that biomarker binarization reduced the accuracy for tau and neurodegeneration predictors, but not Aβ (Figure 2B). Relative to the model with all binary predictors (mean RMSE for A_BIN_/T_BIN_/N_BIN_=0.446), reductions in error were seen when incorporating non-binary categorical tau (A_BIN_/T_CAT_/N_BIN_: RMSE=0.426, t=2.14, p<0.05), continuous tau (A_BIN_/T_CON_/N_BIN_: RMSE=0.424, t=2.42, p<0.05), or continuous neurodegeneration (A_BIN_/T_BIN_/N_CON_: RMSE=0.432, t=2.10, p<0.05) biomarkers. An improvement was also observed when including all continuous biomarkers (A_CON_/T_CON_/N_CON_: RMSE=0.415, t=2.99, p<0.05), but not when including all non-binary biomarkers (A_CAT_/T_CAT_/N_CAT_: RMSE=0.428, t=1.56, p=0.10). The tau SVM (RMSE=0.419, t=2.02, p<0.05) and the AT(N) SVM model (RMSE=0.405, t=3.42, p<0.01) also outperformed the all-binary model. Models which replaced the binary Aβ definition with a non-binary categorical (A_CAT_/T_BIN_/N_BIN_: RMSE=0.446, t=0.26, p=0.60) or continuous (A_CON_/T_BIN_/N_BIN_: RMSE=0.447, t=0.24, p=0.71) version did not improve accuracy. Improvements were also not seen for Aβ and neurodegeneration SVMs (p>0.05).

We found no differences in accuracy between models which used PVC for tau SUVRs and ones with no correction (eFigure 1, all p>0.05).

### Feature importance and model interpretation

For models which applied nested cross-validation to group biomarkers based on AT(N) category and variable type, the best performing predictors were highly consistent across folds, suggesting that some biomarker definitions generally outperformed others measuring the same pathology (Figure 3A). This was particularly true for tau and neurodegeneration models, where the same biomarker definitions were selected in more than 95% of folds. There was slightly more variation for Aβ, but the best performing biomarker was still chosen at least 67% of the time. For PET binarization, previously published cutoffs accounted for 100% of selected Aβ biomarkers, but only 1% of tau biomarkers (the other 99% being a GMM applied to the MTT SUVR). For non-binary categorical measures of PET, staging systems appeared to generally outperform other approaches, appearing in 76% of selected Aβ models and 100% of tau models.

**Figure 3:**
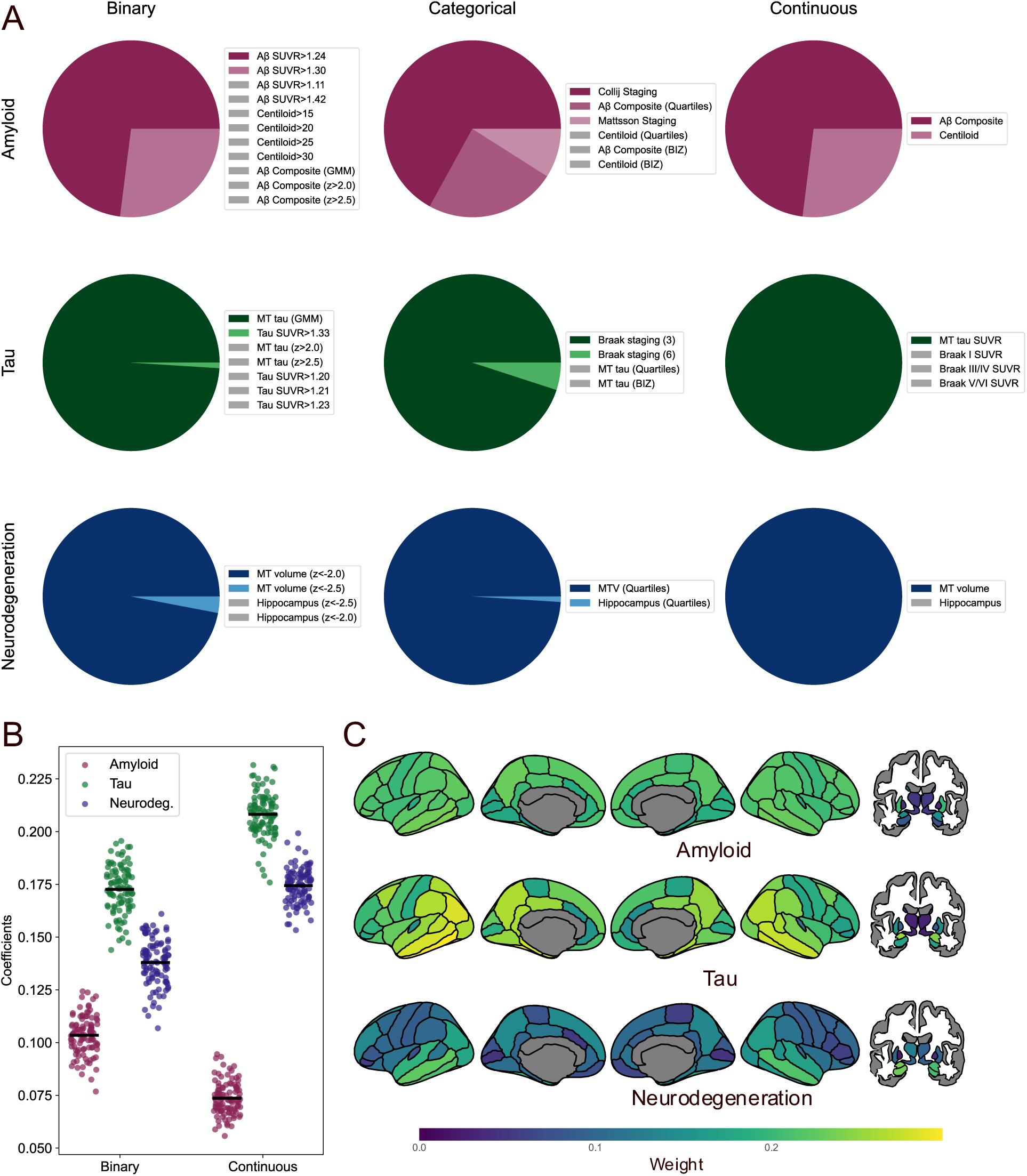
Feature importance analysis. **A.** Pie charts showing which biomarkers were selected as the best performing from cross-validation (100 training fold instances). Biomarkers shown with gray coloring were not selected in any iteration. **B.** Coefficients for the Aβ, tau, and neurodegeneration predictor in cross-validated linear models. Values are taken from 100 instances of the all binary (A_BIN_/T_BIN_/N_BIN_) and all continuous (A_CON_/T_CON_/N_CON_) models. **C.** Brain maps showing average regional feature importance derived from the A_SVM_/T_SVM_/N_SVM_ model.

Inspection of model coefficients highlighted the relative importance of tau for predicting cognition. For linear models which included multiple biomarkers as predictors, coefficients were highest for tau, followed by neurodegeneration and Aβ (Figure 3B). When considering continuous biomarkers in particular, weights for Aβ were much lower than those of tau or neurodegeneration. Similarly, the weights for tau features were on average higher than those of Aβ or atrophy in the multimodal SVM model (Figure 3C). Cortical weights for tau and neurodegeneration highlighted medial and lateral temporal structures, while Aβ weights were more homogenous. Subcortical regions were weighted lower than cortical ones, except for tau uptake and gray matter volume in the amygdala and hippocampus. Similar spatial patterns were observed when considering the SVM weights from separate Aβ, tau, and neurodegeneration models (eFigure 2).

We used our cross-validated modeling to identify the optimal cutoffs for Aβ and tau binarization in our cognitive modeling experiments. Our results indicated SUVR cutoffs of 1.26 (range: 1.24-1.30) for Aβ and 1.44 (range: 1.33-1.45) for tau (Figure 4).

**Figure 4.**
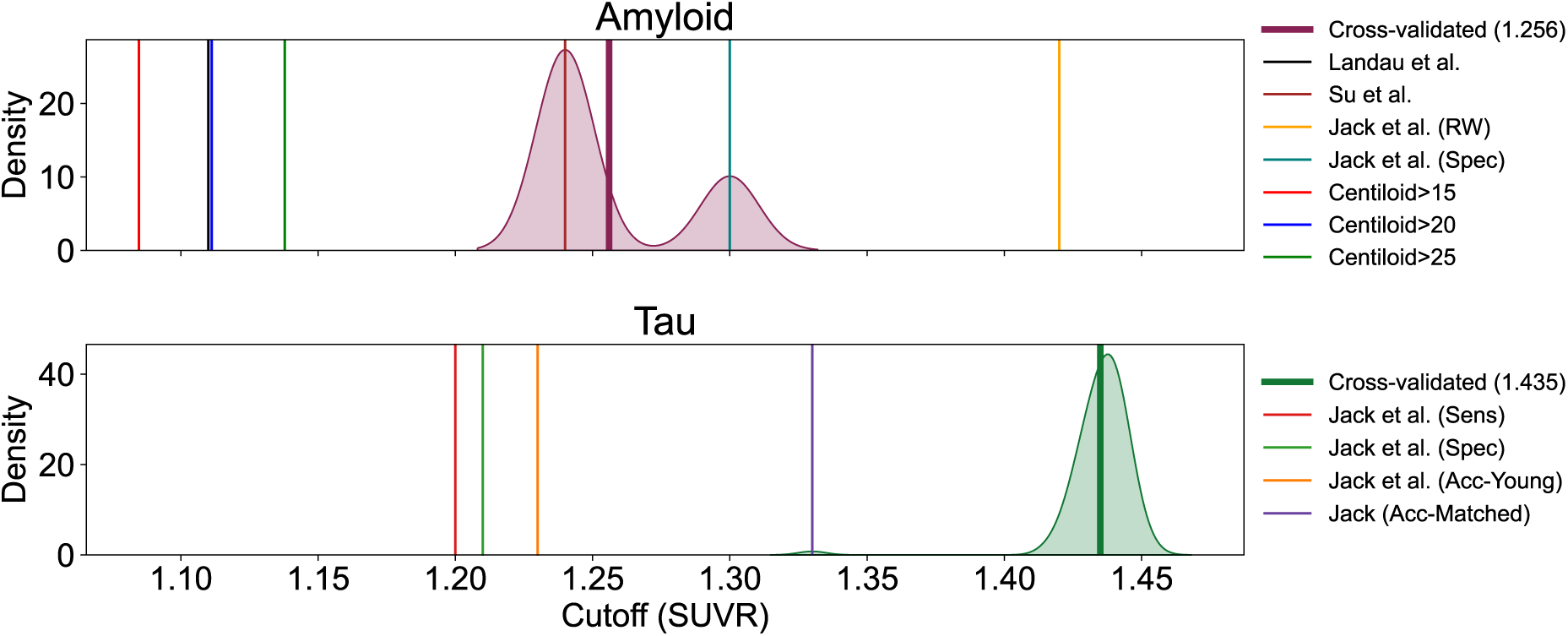
Estimated cutoffs for Aβ-and tau-PET binarization. **A.** The kernel density estimation of selected cutoffs for Aβ (100 cross-validation iterations) is shown in maroon, with the mean value (1.256) highlighted with the bold vertical line. **B.** Same as A., but for tau and shown in green (mean value: 1.435). In both panels, other vertical lines show other pre-defined cutoff values that were tested.

### Cognitive modeling in CU and CI populations

We also trained separate models to optimize the prediction of cognitive scores for individuals who were CU (CDR=0) and those who were CI (CDR>0). Like our results in the whole sample, we found that almost all biomarker models tested resulted in a significant improvement in accuracy relative to a baseline model with just covariates (eFigure 3). These benefits were observed for both CU (range in RMSE reduction: 3.9-15.1%) and CI (range in RMSE reduction: 7.2-30.2%) settings. The only exception was for categorical neurodegeneration models in those CU, where the difference was non-significant (t=1.65, p=0.05). The best performing models in the CU and CI populations were the all-continuous (A_CON_/T_CON_/N_CON_: RMSE=0.349, t=6.1, p<0.001) and AT(N) SVM models (A_SVM_/T_SVM_/N_SVM_: RMSE=0.452, t=7.5, p<0.001), respectively.

Similarly to the whole-sample results, non-binary measures of tau and neurodegeneration, but not Aβ, provided additional accuracy for modeling PHC_Global_ in CU and CI individuals (eFigure 4). Larger benefits were seen for models including non-binary tau and neurodegeneration in CI individuals relative to CU individuals. For CU individuals (mean RMSE for A_BIN_/T_BIN_/N_BIN_: 0.381), we observed improvements when including non-binary categorical tau (A_BIN_/T_CAT_/N_BIN_: RMSE=0.366, t=2.8, p<0.05), continuous tau (A_BIN_/T_CON_/N_BIN_: RMSE=0.366, t=2.6, p<0.05), or continuous biomarkers (A_CON_/T_CON_/N_CON_: RMSE=0.359, t=2.5, p<0.05). Considering CI people (mean RMSE for A_BIN_/T_BIN_/N_BIN_: 0.523), we observed increases in accuracy for the categorical tau (A_BIN_/T_CAT_/N_BIN_: RMSE=0.490, t=2.6, p<0.05), continuous tau (A_BIN_/T_CON_/N_BIN_: RMSE=0.485, t=3.3, p<0.01), continuous neurodegeneration (A_BIN_/T_BIN_/N_CON_: RMSE=0.495, t=3.4, p<0.01), continuous AT(N) (A_CON_/T_CON_/N_CON_: RMSE=0.479, t=3.4, p<0.01), tau SVM (T_SVM_: RMSE=0.484, t=2.4, p<0.05), and AT(N) SVM (A_SVM_/T_SVM_/N_SVM_: RMSE=0.442, t=4.8, p<0.001) models.

### Modeling longitudinal cognition

The pattern of results we observed were largely consistent when modeling prospective change in cognition. All model varieties tested were significantly more accurate for predicting longitudinal change in PHC_Global_ (range in RMSE reduction: 4.1-30.3%, all p<0.05) relative to a covariate-only model (eFigure 5a), with the largest benefits seen for the all-continuous (A_CON_/T_CON_/N_CON_: RMSE=0.386, t=5.83, p<0.001), multimodal SVM (A_SVM_/T_SVM_/N_SVM_: RMSE=0.368, t=5.44, p<0.001), and tau SVM (T_SVM_: RMSE=0.373, t=5.44, p<0.001). No biomarker linear models with non-binary measures improved prediction accuracy relative to an all-binary baseline (eFigure 5b, all p>0.05). However, the tau SVM (T_SVM_: RMSE=0.373, t=3.05, p<0.01) and multimodal SVM (A_SVM_/T_SVM_/N_SVM_: RMSE=0.368, t=3.54, p<0.01) were still significantly more accurate at predicting change in PHC_Global_ than a model consisting of all-binary predictors.

**Figure 5.**
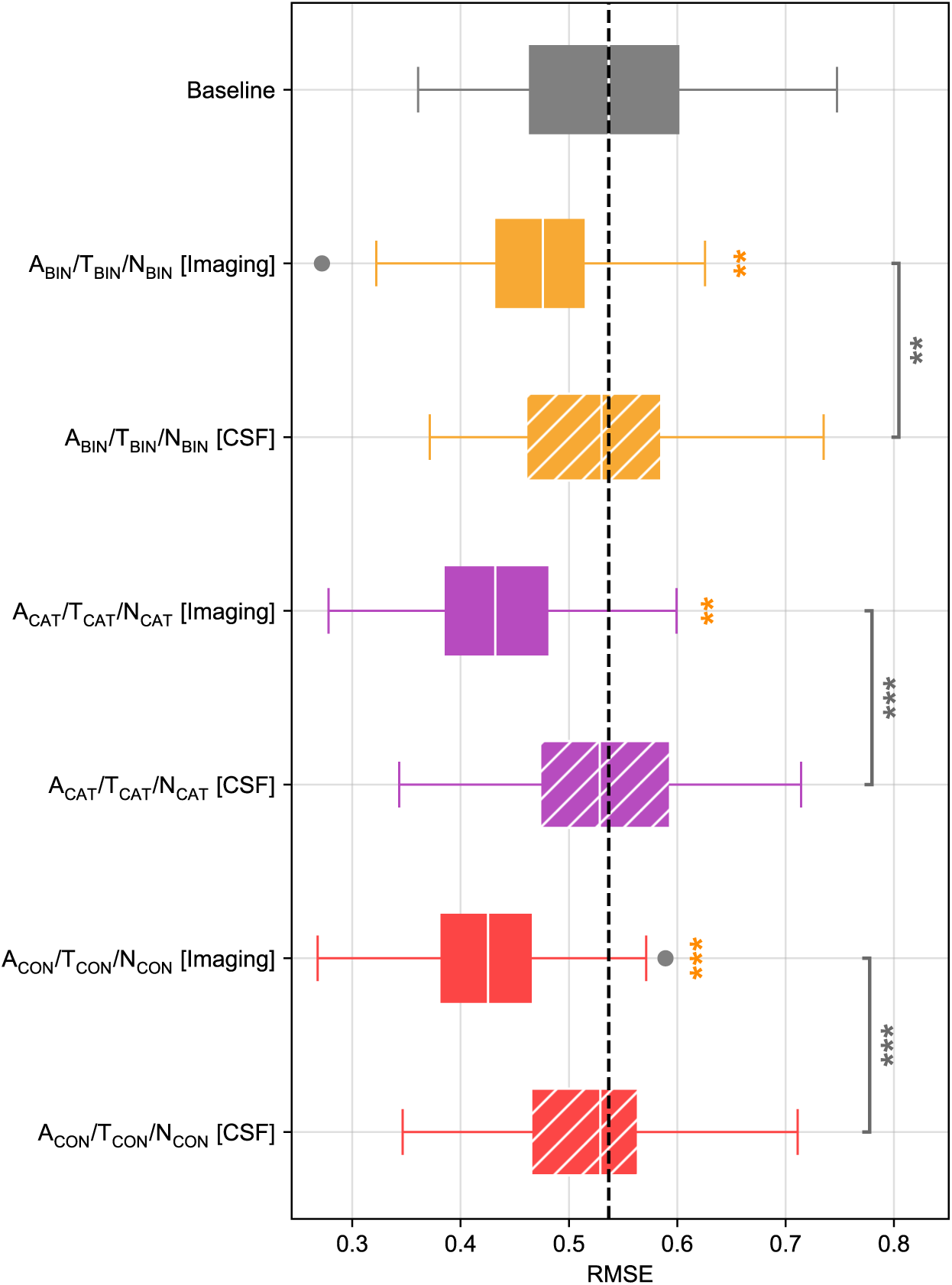
Boxplots comparing accuracy cognition predictions using image-based (solid) and CSF-based (hatched) biomarkers. Values are RMSEs taken from 100 cross-validation instances. Colors are used to indicated the variable type of included biomarkers (yellow: binary, purple: non-binary categorical, red: continuous). Gold stars indicate a significant improvement in accuracy relative to the baseline model (only covariates). Gray stars and bars highlight significant pairwise differences between individual models. Statistical results are derived from Nadeau-Bengio t-tests with correction for multiple comparisons (*p<0.05, **p<0.01, ***p<0.001).

### Modeling of individual neuropsychological domains

We also observed similar patterns of accuracy differences for non-binary biomarker definitions when modeling neuropsychological domains instead of PHC_Global_. Significant benefits were only observed for models which included non-binary or SVM-based assessments of tau and neurodegenerative pathology (eFigure 6). Models which included non-binary definitions of Aβ alone did not surpass the all-binary model for any neuropsychological domain. Continuous tau and neurodegeneration measures improved accuracy for prediction of PHC_EF_ (p<0.05) and PHC_Visual_ (p<0.05), while non-binary categorical measures only improved prediction of PHC_Visual_ (p<0.05). The AT(N) SVM had significantly higher accuracy for modeling PHC_EF_ (p<0.01), PHC_Visual_ (p<0.05), and PHC_Memory_ (p<0.05). No significant differences were observed for PHC_Language_ prediction accuracy (all p>0.05).

### Comparison of image-based and CSF-based models

We observed that CSF-based models performed relatively poorly for modeling cognition. Models which incorporated CSF-based biomarkers, as opposed to imaging-based ones, did not perform better than a baseline model consisting of only covariates (Figure 5 & eFigure 7, all p>0.05). Moreover, imaging-based models were significantly more accurate than CSF-based models. This was true for binary (t=2.81, p<0.01), non-binary categorical (t=3.68, p<0.01), and continuous (t=3.96, p<0.01) biomarker definitions.

## Discussion

AD biomarkers differ from each other along various axes such as the underlying pathology they measure (e.g. Aβ, tau), the modality (e.g., imaging, CSF, blood), and measurement characteristics (e.g., variable type). Our analyses indicate that differences along these dimensions result in considerable variability when imaging biomarkers are utilized in downstream tasks. We show that even biomarkers which assess the same pathology exhibit a range in accuracy when applied for modeling cognition. Additionally, we demonstrate that multivariate machine learning approaches can surpass traditional biomarker definitions for cognitive prediction in AD. Careful consideration should be applied when selecting biomarker definitions for predictive tasks, as certain operationalizations may be relatively less informative than other variants. While we specifically focus on cognitive prediction, our results may be relevant for other settings where researchers wish to quantify AD pathology.

While nearly all tested biomarkers provided predictive gains when modeling cognition, some categories of biomarkers yielded consistently larger improvements than others. Multiple analyses demonstrated that tau biomarkers exhibited stronger associations with cognition than assessments of Aβ or neurodegeneration, a well-documented finding.^42–44^ Feature importance analyses indicated that tau predictors were weighted higher than Aβ or neurodegeneration measures in models which incorporated all three AT(N) categories. However, combined AT(N) models generally outperformed unimodal ones, even when tau was the single biomarker included. Thus, while measures of tau are important indicators of cognitive decline, incorporation of measures spanning other pathologies is warranted for enhancing predictive accuracy.

We observed that SVM models outperformed more traditional linear models of cognition. Tau and multimodal SVMs were the best predictors overall in many experiments. These models were also the only models which outperformed binary biomarker models for prediction of longitudinal cognitive decline. Aβ and neurodegeneration SVMs were also relatively stronger than other individual biomarker definitions of these pathologies. The superior performance of SVM models suggest that there may be key predictive signal occurring in brain regions external to the manually defined meta-ROIs which are utilized in most of the biomarker definitions we tested. However, the SVMs also allowed for non-linear transformations of input features, making them relatively more powerful models.

Inclusion of non-binary tau and neurodegeneration predictors led to small but consistent improvements in accuracy relative to binary alternatives. However, binary Aβ measures performed equally to non-binary ones. As such, our findings indicate that binarization along dimensions of tau and neurodegeneration (e.g., labeling individuals as T+/-or N+/-) may obfuscate information relevant to the prediction of cognitive decline. On the other hand, dichotomization of Aβ status may be sufficient. These notion agrees with revised criteria for diagnosis and staging of AD^2^: their proposed PET staging system includes binary assessment of Aβ (i.e., has AD or not) and multi-level staging of tau based on the extent of progression outside the medial temporal lobe. Interestingly, the specific benefits for tau and neurodegeneration (and not Aβ) were consistent when considering only CU or CI individuals and when modeling some neuropsychological domains (executive functioning and visuospatial performance). These results agree with a previous study which found similar non-dichotomized tau and neurodegeneration for modeling longitudinal cognitive decline^13^. However, we did not replicate their findings showing accuracy improvements when modeling prospective cognition in CU individuals and including non-binary measures of Aβ.

While nearly all the imaging biomarkers we tested improved prediction of cognitive impairment, the same was not true for CSF counterparts. Models which incorporated CSF biomarkers as predictors did not perform better than baseline models which only included standard covariates, regardless of the analyte or its operationalization. Previous findings have similarly demonstrated stronger associations for imaging biomarkers and cognitive scores in AD, relative to fluid biomarkers^45^. Importantly, the CSF analytes tested largely reflect earlier pathological cascades which likely develop and saturate prior to the onset of neurodegeneration and cognitive decline^2,46^. As such, they may be less suited for providing direct associations with cognitive decline, and more suited for diagnosis or prediction of future decline.

Our study has limitations which should be considered. First, while we performed a comprehensive and rigorous evaluation of multiple biomarkers, biomarkers such as fluorodeoxyglucose-PET, cortical thickness, and functional imaging were not included in this study. Future studies are warranted to examine them. Second, this study relied only on ADNI because inclusion of other sources posed issues of harmonization and biomarker availability. While ADNI is one of few databases which can enable the analyses we conducted, it is also relatively limited in its inclusion of demographic diversity^47^. As such, our results warrant replication in other datasets.

The growth of large data initiatives has led to an explosion of approaches for biomarker assessment of AD. While picking from the myriads of methods for quantification of AD pathology, it is important for researchers to mind the biological, statistical, and practical characteristics of each approach. Our results demonstrate that different operationalizations of the same pathology can result in variable performance for downstream predictive tasks. More complex indices of pathology may be superior to dichotomous alternatives, particularly for measures of neurodegeneration and tau. Finally, data-driven, machine learning approaches may be preferable for identifying biomarker contributions to cognitive decline.

## Data Availability

All data used in this study are accessible from ADNI following formal data usage agreements (https://adni.loni.usc.edu/). All code for this project will be shared at the following repository: https://github.com/sotiraslab/earnest_ad_biomarker_modeling.

https://github.com/sotiraslab/earnest_ad_biomarker_modeling

## Acknowledgement

The authors thank the staff for the Washington University Center for High Performance Computing who helped enable this work. Computations were performed using the facilities of the Washington University Research Computing and Informatics Facility (RCIF). The RCIF has received funding from NIH S10 program grants: 1S10OD025200-01A1 and 1S10OD030477-01.

