## Supplementary Material for "Comprehensive evaluation of AT(N) imaging biomarkers for predicting cognition"

### Supplement

### eMethods

#### PET-based staging systems

PET-based staging systems were included as non-binary categorical measures of Aβ and tau pathology. We included two systems for Aβ: a three-stage system developed by Mattsson et al[1] and a four-stage system developed by Collij et al[2]. We included Braak staging for tau[3,4], including both a three-stage and six-stage version. Each stage in each system was associated with a collection of Freesurfer ROIs (stage composite), as shown in eTable 2, with advancing stages indicating regions where pathology spreads over the disease course.

The method for Mattsson staging and Braak staging were similar and proceeded as follows. First, an average SUVR was calculated in each stage composite using a volume weighted average of bilateral regions. Next, Gaussian mixture models (GMM) were fit to the distribution of uptake in each stage composite to estimate a binary cutoff for stage positivity (as described in the main text Methods: Image-based biomarker definitions). These cutoffs allowed us to assign binary positivity measures for each subject in each stage composite. Disease stages were then assigned based on individual patterns of positivity: to be assigned a given disease stage, an individual had to exhibit positivity for that stage and all prior ones. In case an individual was positive for a given stage but not all prior stages, they were marked as non-stageable. Individuals could also be assigned stage 0 if they were not positive for pathology in any stage composite.

Collij staging was slightly different in that positivity for a stage composite was based on being positive for pathology in most ROIs corresponding to a given stage. That is, GMMs were fit to each Freesurfer ROI individually, and positivity for a stage composite was defined by exhibiting supra-threshold uptake in 50% or more of the associated ROIs. Disease stage assignment then proceeded in the same manner as for Mattsson and Braak staging (individuals needed to be positive for a given disease stage and all prior stages).

#### Cross-validation experiments

Cross-validated modeling was implemented in Python (v3.10) using scikit-learn (v1.4.2). For all cross-validation experiments, CDR status was used as a stratifying variable. Random samples were also seeded, such that the individuals of each testing fold were the same across experiments with the same input data. All cross-validation experiments had 10 outer folds and were repeated 10 times to generate 100 out of sample error estimates for each tested model.

We first ran models for each biomarker separately to assess the predictive value of each included definition. These experiments used a non-nested, 10-fold cross-validation. In each iteration, training data were used to fit linear regressions where $PHC_{Global}$ was predicted from a single biomarker and covariates (age, sex, *APOE* E4 positivity). That is, separate models were fit for each biomarker. Biomarker definitions with tunable parameters (Z-scores, GMMs) were also fit with training data. A baseline model was also trained in each iteration which only included covariates as predictors. Trained models were then evaluated on the testing fold data, and the prediction accuracies for each were calculated as root mean squared error (RMSE).

We next created combinatorial models to test if combination of biomarkers improved prediction accuracy and if models incorporating continuous or non-categorical binary biomarkers outperformed models with binary biomarkers. For these experiments, we used nested cross-validation with 10 outer folds and 5 inner folds. In the inner cross-validation, a model selection procedure was applied to identify the best performing individual predictors (like the non-nested cross-validation experiment described above). Inner training data were used to fit linear models predicting $PHC_{Global}$ with single biomarkers plus covariates as predictors. Inner testing data were used to measure the out-of-sample accuracy of these models. We then grouped the models based on the pathology (AT(N)) and variable type (binary/non-binary categorical/continuous) and selected the best performing biomarker definitions (lowest average RMSE across 5 test folds). The outer cross-validation was then used for training new linear models which combined the biomarkers selected from the inner cross-validation. eTable 3 shows all the combinatorial linear models that were evaluated in the outer loop using this training scheme.

We also used support vector machine (SVM) regression to directly predict $PHC_{Global}$ from imaging data. SVMs were trained using regional Aβ uptakes, tau uptakes, or gray matter volumes as input features. An additional model combined all these regional imaging features into a multimodal predictive model. SVM training used a nested cross-validation design with 10 outer folds and 5 inner folds. The inner loop was used for hyperparameter tuning for the regularization parameter $C$, the kernel, and the kernel coefficient $\gamma$. Search spaces were informed by consensus recommendations[5] and from pilot experiments: kernel (linear or radial basis function [RBF]), $C$ with linear kernel $(2^{-10}, 2^{-9}, \cdots, 2^{-1}, 2^{0})$, $C$ with RBF kernel $(2^{-5}, 2^{-3}, \cdots, 2^{13}, 2^{15})$, $\gamma$ with RBF kernel $(2^{-15}, 2^{-13}, \cdots, 2^{1}, 2^{3})$. The best hyperparameters were determined from the inner loop (lowest average RMSE across 5 test folds) and used to retrain and evaluate SVM models in the outer loop.

#### Feature importance analyses

We ran additional post-hoc analyses to probe feature importance for our cross-validated linear modeling. For models which applied a model selection to filter biomarker definitions, we created pie charts showing the specific biomarker definitions which were selected as the best performing over repeated cross-validation iterations. We also extracted and plotted the (standardized) linear model coefficients for the Aβ, tau, and neurodegeneration biomarkers in all binary and all continuous models (non-binary categorical models were omitted because coefficient interpretation is less straightforward for non-binary categorical features). Finally, we visualized the cutoff values that were selected for Aβ and tau from models with all binary definitions.

For SVM models, we visualized the feature importance of individual brain regions for Aβ, tau, and neurodegeneration features. Following previous work[6,7], feature importance values were generated by calculating the covariance of each feature and $PHC_{Global}$. We generated brain maps showing the average feature importance across 100 out-of-sample model predictions. Maps were generated for the combined SVM and for each unimodal SVM.

### eTables

| Name | Pathology | Variable type | Description |
|---|---|---|---|
| Aβ composite | Aβ | Continuous | Aβ SUVR in summary composite region |
| Centiloid | Aβ | Continuous | Linear transformation of Aβ composite SUVR[8] |
| Aβ SUVR>1.11 | Aβ | Binary | Cutoff from Landau et al.[9] |
| Aβ SUVR>1.24 | Aβ | Binary | Cutoff from Su et al.[10] |
| Aβ SUVR>1.42 | Aβ | Binary | Cutoff from Jack et al.[11] (reliable worsening) |
| Aβ SUVR>1.30 | Aβ | Binary | Cutoff from Jack et al.[11] (specificity) |
| Centiloid>15 | Aβ | Binary | Binary cutoff for Centiloid |
| Centiloid>20 | Aβ | Binary | Binary cutoff for Centiloid |
| Centiloid>25 | Aβ | Binary | Binary cutoff for Centiloid |
| Centiloid>30 | Aβ | Binary | Binary cutoff for Centiloid |
| Aβ composite (GMM) | Aβ | Binary | Aβ composite SUVR binarized with a GMM |
| Aβ composite (z>2.0) | Aβ | Binary | Z-score cutoff of 2 for Aβ composite SUVR |
| Aβ composite (z>2.5) | Aβ | Binary | Z-score cutoff of 2.5 for Aβ composite SUVR |
| Aβ composite (Quartiles) | Aβ | Non-binary categorical | Quartiles of the Aβ composite SUVR |
| Centiloid (Quartiles) | Aβ | Non-binary categorical | Quartiles of Centiloid |
| Mattsson staging | Aβ | Non-binary categorical | Aβ –PET staging system[2] |
| Collij staging | Aβ | Non-binary categorical | Aβ –PET staging system[1] |
| Aβ composite (BIZ) | Aβ | Non-binary categorical | Binarization with an intermediate zone for Aβ composite |
| Centiloid (BIZ) | Aβ | Non-binary categorical | Binarization with an intermediate zone for Centiloid |
| MT tau SUVR | Tau | Continuous | Tau SUVR in meta-temporal composite region |
| Braak I SUVR | Tau | Continuous | Tau SUVR in Braak I composite region |
| Braak III/IV SUVR | Tau | Continuous | Tau SUVR in Braak III/IV composite region |
| Braak V/VI SUVR | Tau | Continuous | Tau SUVR in Braak V/VI composite region |
| MT tau (GMM) | Tau | Binary | Meta-temporal tau SUVR binarized with a GMM |
| MT tau (z>2.0) | Tau | Binary | Z-score cutoff of 2 for MT tau SUVR |
| MT tau (z>2.5) | Tau | Binary | Z-score cutoff of 2.5 for MT tau SUVR |
| Tau SUVR>1.20 | Tau | Binary | Cutoff from Jack et al.[11] (sensitivity) |
| Tau SUVR>1.21 | Tau | Binary | Cutoff from Jack et al.[11] (specificity) |
| Tau SUVR>1.23 | Tau | Binary | Cutoff from Jack et al.[11] (accuracy-young) |
| Tau SUVR>1.33 | Tau | Binary | Cutoff from Jack et al.[11] (accuracy-matched) |
| MT tau (Quartiles) | Tau | Non-binary categorical | Quartiles of MT tau SUVR |
| MT tau (BIZ) | Tau | Non-binary categorical | Binarization with an intermediate zone for MT tau SUVR |

| | | | |
|---|---|---|---|
| Braak staging (3) | Tau | Non-binary categorical | Braak staging based on 3 stage model (I, III/IV, V/VI) |
| Braak staging (3) | Tau | Non-binary categorical | Braak staging based on 6 stage model (I, III, V, V, VI) |
| Hippocampus | Neurodegen. | Continuous | Hippocampal volume |
| MT volume | Neurodegen. | Continuous | Volume of the MT composite region |
| Hippocampus (z<-2.0) | Neurodegen. | Binary | Z-score cutoff of 2.0 for hippocampal volume |
| Hippocampus (z<-2.5) | Neurodegen. | Binary | Z-score cutoff of 2.5 for hippocampal volume |
| MT volume (z<-2.0) | Neurodegen. | Binary | Z-score cutoff of 2.0 for MT volume |
| MT volume (z<-2.5) | Neurodegen. | Binary | Z-score cutoff of 2.5 for MT volume |
| Hippocampus (Quartiles) | Neurodegen. | Non-binary categorical | Quartiles of hippocampal volume |
| MT volume (Quartiles) | Neurodegen. | Non-binary categorical | Quartiles of MT volume |

**eTable 1.** Listing of all image-based AT(N) biomarkers used in cognitive prediction models.

| Name | Citation | Regions |
|---|---|---|
| Aβ composite | [12,13] | caudal middle frontal, lateral orbitofrontal, medial orbitofrontal, pars opercularis, pars orbitalis, pars triangularis, rostral middle frontal, superior frontal, frontal pole, caudal anterior cingulate, isthmus cingulate, posterior cingulate, rostral anterior cingulate, inferior parietal, precuneus, superior parietal, supramarginal, inferior temporal, middle temporal, superior temporal |
| MT | [11] | entorhinal, amygdala, fusiform, inferior temporal, middle temporal |
| Braak I | [14,15] | entorhinal |
| Braak III | | parahippocampal, fusiform, lingual, amygdala |
| Braak IV | | middle temporal, caudal anterior cingulate, rostral anterior cingulate, posterior cingulate, isthmus cingulate, insula, inferior temporal, temporal pole |
| Braak V | | superior frontal, lateral orbitofrontal, medial orbitofrontal, frontal pole, caudal middle frontal, rostral middle frontal, pars opercularis, pars orbitalis, pars triangularis, lateral occipital, supramarginal, inferior parietal, superior temporal, superior parietal, precuneus, bank of the superior temporal sulcus, transverse temporal |
| Braak VI | | pericalcarine, postcentral, cuneus, precentral, paracentral |
| Mattsson Early | [1] | precuneus, posterior cingulate, isthmus cingulate, insula, medial orbitofrontal, lateral orbitofrontal |
| Mattsson Intermediate | | bank SSTS, caudal middle frontal, cuneus, frontal pole, fusiform, inferior parietal, inferior temporal, lateral occipital, middle temporal, parahippocampal, pars opercularis, pas orbitalis, pars triangularis, putamen, rostral anterior cingulate, rostral middle frontal, supramarginal |
| Mattsson Late | | lingual, pericalcarine, paracentral, precentral, postcentral |
| Colllij 1 | [2] | posterior cingulate, isthmus cingulate, anterior cingulate (caudal+rostral) |
| Collij 2 | | lateral orbitofrontal, paracentral, precuneus, medial orbitofrontal, inferior frontal (pars opercularis+pas orbitalis+pars triangularis) |
| Collij 3 | | insula, fusiform, precentral, inferior temporal, parahippocampal, collijinferiorfrontal, superior frontal, lingual, supramarginal, inferior parietal, cuneus, middle frontal (rostral+caudal) |
| Collij 4 | | lateral occipital, superior parietal, middle temporal, superior temporal, postcentral, entorhinal, frontal pole, temporal pole |

**eTable 2.** List of Freesurfer regions used for composite regions. Citations are included for papers which defined these composites. For Collij stging composites, regions shown joined by plus signs are meta-ROIs constructed prior to averaging for the composite, as described in the original paper[2]. Aβ=amyloid-beta, MT=meta-temporal, ROI=region of interest.

| Model | Aβ | Tau | Neurodegen |
| --- | --- | --- | --- |
| Baseline | - | - | - |
| $A_{BIN}$ | Binary | - | - |
| $T_{BIN}$ | - | Binary | - |
| $N_{BIN}$ | - | - | Binary |
| $A_{CAT}$ | Non-binary categorical | - | - |
| $T_{CAT}$ | - | Non-binary categorical | - |
| $N_{CAT}$ | - | - | Non-binary categorical |
| $A_{CON}$ | Continuous | - | - |
| $T_{CON}$ | - | Continuous | - |
| $N_{CON}$ | - | - | Continuous |
| $A_{BIN}/T_{BIN}/N_{BIN}$ | Binary | Binary | Binary |
| $A_{CAT}/T_{CAT}/N_{CAT}$ | Non-binary categorical | Non-binary categorical | Non-binary categorical |
| $A_{CON}/T_{CON}/N_{CON}$ | Continuous | Continuous | Continuous |
| $A_{CAT}/T_{BIN}/N_{BIN}$ | Non-binary categorical | Binary | Binary |
| $A_{BIN}/T_{CAT}/N_{BIN}$ | Binary | Non-binary categorical | Binary |
| $A_{BIN}/T_{BIN}/N_{CAT}$ | Binary | Binary | Non-binary categorical |
| $A_{CON}/T_{BIN}/N_{BIN}$ | Continuous | Binary | Binary |
| $A_{BIN}/T_{CON}/N_{BIN}$ | Binary | Continuous | Binary |
| $A_{BIN}/T_{BIN}/N_{CON}$ | Binary | Binary | Continuous |

**eTable 3.** List of linear models predicting $PHC_{Global}$ from AT(N) biomarkers. Models were trained with a nested-cross validation scheme where the inner loop was used to select the best performing biomarkers, after grouping by pathology (Aβ, tau, neurodegeneration) and variable type. BIN, CAT, and CON are used to represent binary, non-binary categorical, and continuous variables (respectively). Dashes are shown to indicate omission of the corresponding biomarker in the model. All models also had age, sex, and *APOE* E4 status included as covariates.

| | CDR=0.0 | CDR=0.5 | CDR=1.0+ | p-value |
|---|---|---|---|---|
| **n** | 223 | 130 | 30 | |
| **Age** | 73.76 (7.07) | 75.73 (8.32) | 77.78 (8.48) | **0.005** |
| **Sex (M/F)** | 96/127 | 75/55 | 16/14 | **0.026** |
| ***APOE* E4+** | 79 (35.4%) | 45 (34.6%) | 13 (43.3%) | 0.659 |
| **Centiloid** | 19.92 (35.65) | 41.74 (54.70) | 69.79 (48.77) | **<0.001** |
| **$PHC_{Global}$** | 0.84 (0.36) | 0.35 (0.46) | -0.47 (0.55) | **<0.001** |

**eTable 4:** Characteristics for the subsample with longitudinal cognitive followup.

| | CDR=0.0 | CDR=0.5 | CDR=1.0+ | p-value |
|---|---|---|---|---|
| **n** | 143 | 83 | 20 | |
| **Age** | 73.29 (7.50) | 74.26 (8.31) | 75.91 (9.34) | 0.322 |
| **Sex (M/F)** | 59/94 | 54/29 | 10/10 | **0.003** |
| ***APOE* E4+** | 49 (34.3%) | 29 (34.9%) | 12 (60.0%) | 0.076 |
| **Centiloid** | 19.94 (38.52) | 45.26 (55.40) | 74.68 (48.25) | **<0.001** |
| **$PHC_{Global}$** | 0.87 (0.35) | 0.32 (0.50) | -0.27 (0.53) | **<0.001** |

**eTable 5:** Characteristics for the subsample with imaging and CSF biomarker assessments.

### eFigures

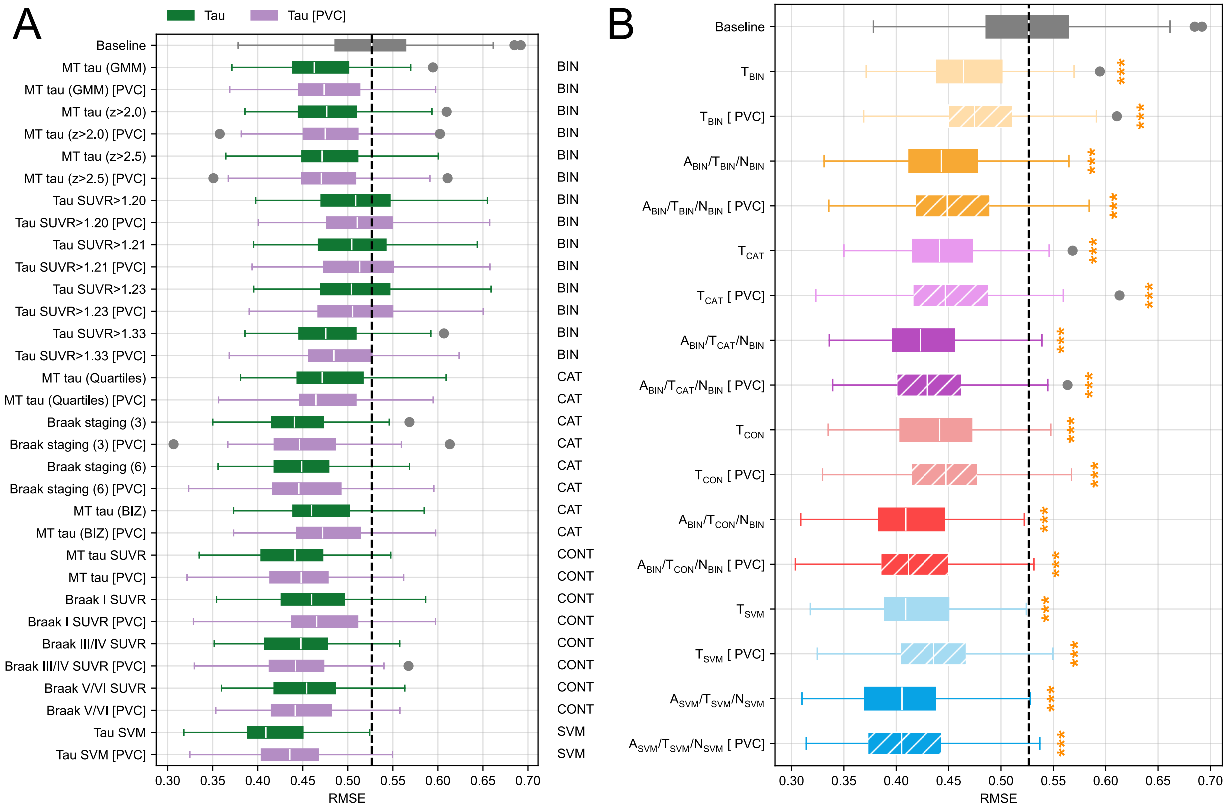


**eFigure 1:** Comparison of partial volume corrected (PVC) and non-PVC tau predictors for modeling cognition. **A.** Boxplots of cross-validated accuracy (RMSE) for models including a single tau predictor and covariates. **B.** Boxplots of cross-validated accuracy (RMSE) for models with the best selected tau predictors. Solid colors indicate models without tau PVC, while hatches indicate models with tau PVC. In both panels, the baseline model (just covariates) is shown in gray, with the dotted line indicating its mean performance. Gold stars indicate a significant improvement in accuracy relative to the baseline model (*p<0.05, **p<0.01, ***p<0.001). No significant differences were found for comparisons of PVC and non-PVC models (all p>0.05).

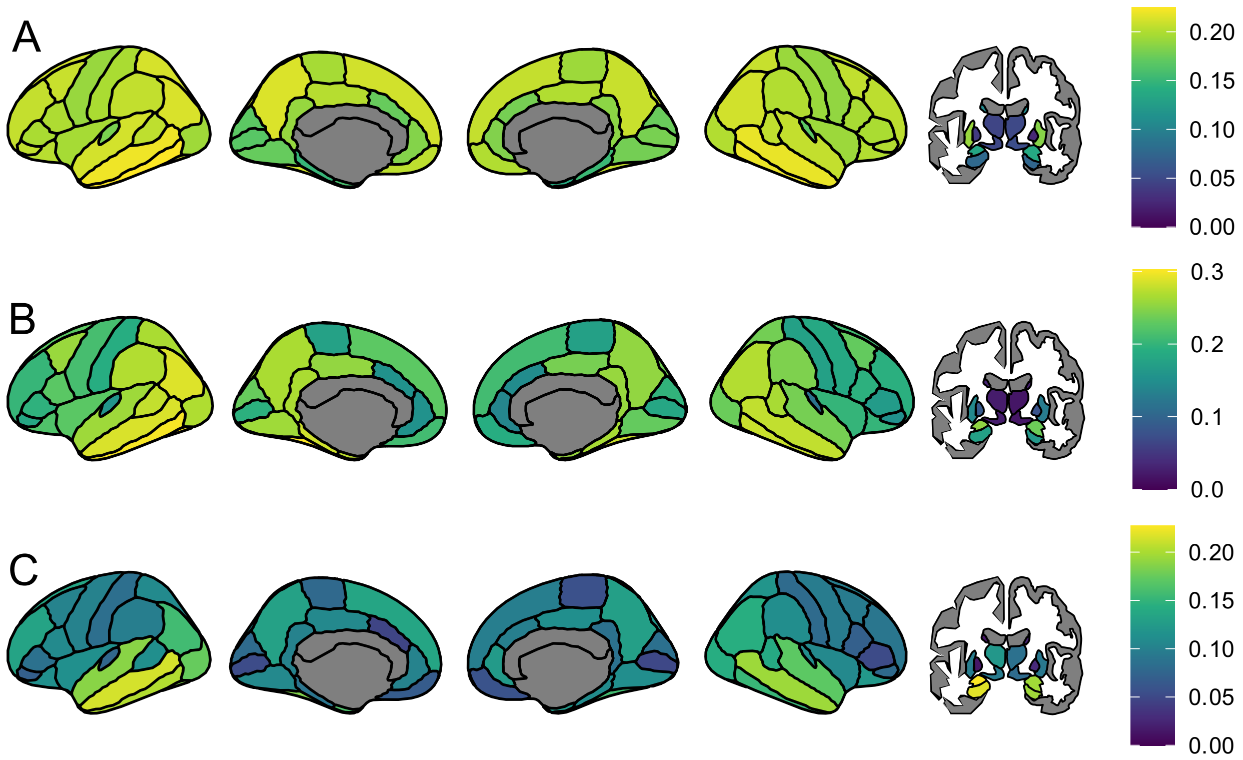


**eFigure 2:** Regional weights for SVM models which included only Aβ (**A,** $A_{SVM}$), tau (**B,** $T_{SVM}$), or gray matter volume (**C,** $N_{SVM}$).

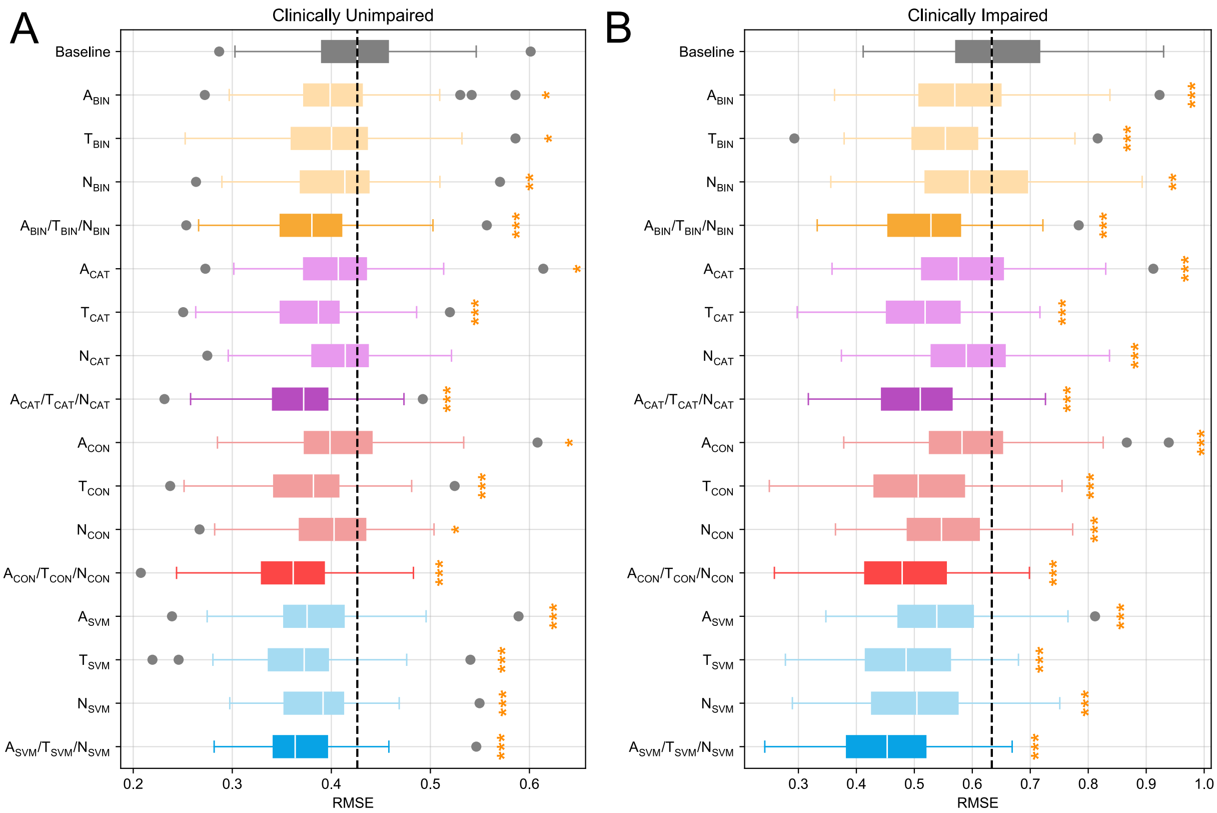


**eFigure 3**: Boxplots showing cross-validated accuracy (RMSE) measures for predicting $PHC_{Global}$ in CU (**A**) and CI (**B**) individuals. Individual and combination biomarker models are compared against a baseline model using only covariates (mean performance indicated by dotted line) to predict $PHC_{Global}$. Colors are used to indicate the variable type of included biomarkers (yellow: binary, purple: non-binary categorical, red: continuous, blue: SVM). Lighter coloring indicates models which only have a single pathology assessment, while darker coloring indicates models which have Aβ, tau, and neurodegeneration biomarkers. Gold stars indicate a significant improvement in accuracy relative to the baseline model (*p<0.05, **p<0.01, ***p<0.001).

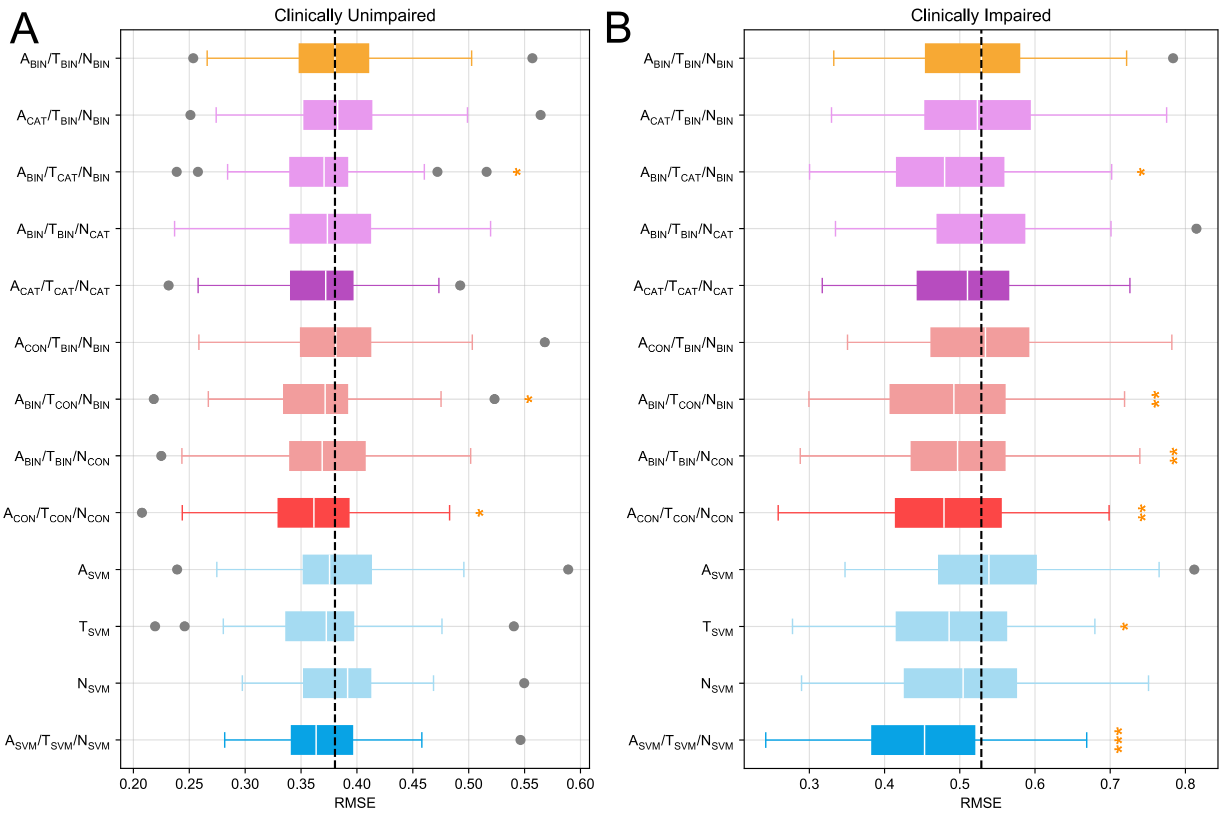


**eFigure 4**: Boxplots showing cross-validated accuracy (RMSE) measures for predicting $PHC_{Global}$ in CU (**A**) and CI (**B**) individuals. Combination biomarker models with non-binary variable types are compared against a baseline model with binary biomarker definitions (mean performance indicated by dotted line). Colors are used to indicated the variable type of included biomarkers (yellow: binary, purple: non-binary categorical, red: continuous, blue: SVM). Lighter coloring indicates models which only have a single pathology assessment, while darker coloring indicates models which have Aβ, tau, and neurodegeneration biomarkers. Gold stars indicate a significant improvement in accuracy relative to the baseline model (*p<0.05, **p<0.01, ***p<0.001).

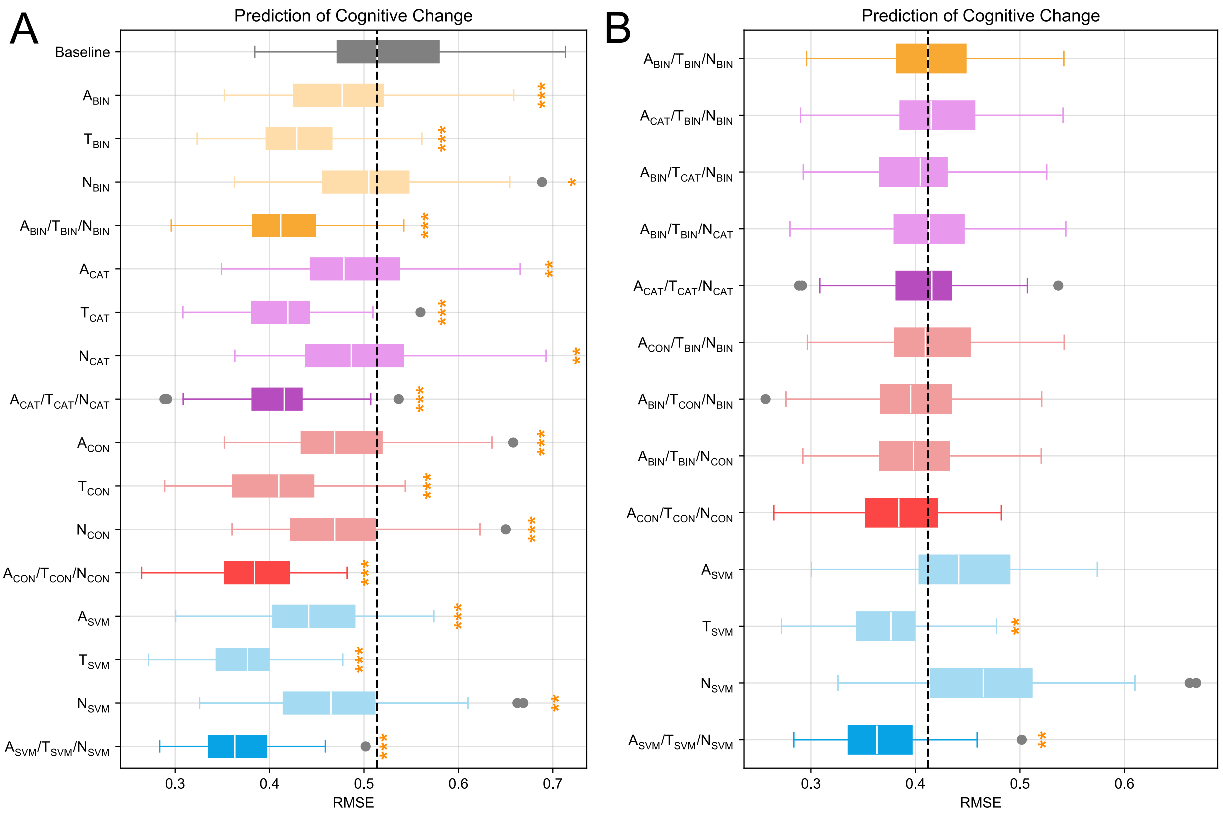


**eFigure 5**: Results from experiments predicting the longitudinal change in $PHC_{Global}$. **A.** Individual and combination biomarker models are compared against a baseline model using only covariates (mean performance indicated by dotted line). **B.** Combination biomarker models with non-binary variable types are compared against a baseline model with binary biomarker definitions (mean performance indicated by dotted line). In both panels, colors are used to indicate the variable type of included biomarkers (yellow: binary, purple: non-binary categorical, red: continuous, blue: SVM). Lighter coloring indicates models which only have a single pathology assessment, while darker coloring indicates models which have Aβ, tau, and neurodegeneration biomarkers. Gold stars indicate a significant improvement in accuracy relative to the topmost model. Gray stars and bars highlight significant pairwise differences between individual models. Statistical results are derived from Nadeau-Bengio t-tests with correction for multiple comparisons (*p<0.05, **p<0.01, ***p<0.001).

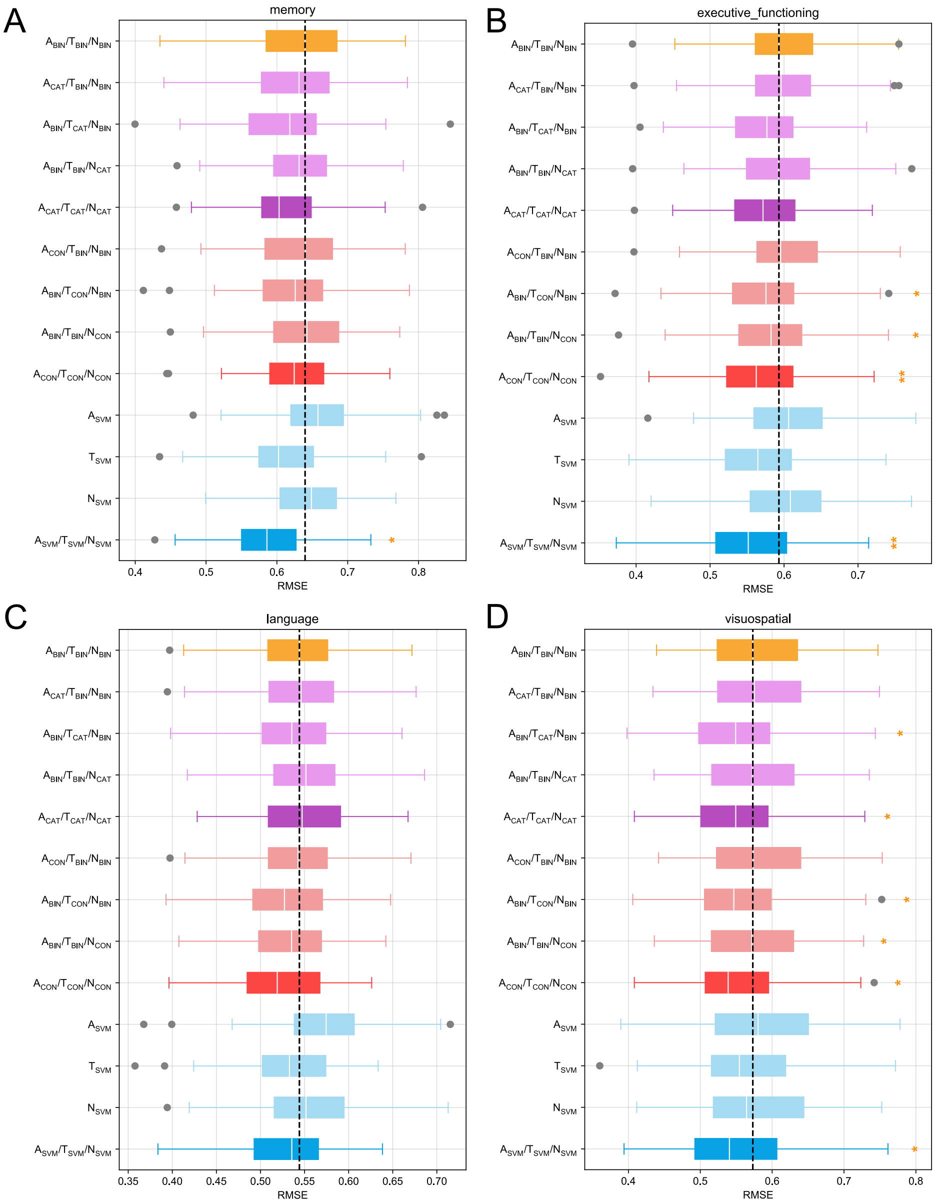


**eFigure 6**: Boxplots showing cross-validated accuracy estimates (RMSE) for models predicting neuropsychological performance from biomarkers. Panels show experiments using memory (A), executive functioning (B), language (C), and visuospatial (D) composites from the PHC as dependent variables. Combination biomarker models with non-binary variable types are compared against a baseline model with binary biomarker definitions (mean performance indicated by dotted line). In all panels, colors are used to indicate the variable type of included ATN biomarkers (yellow: binary, purple: non-binary categorical, red: continuous, blue: SVM). Lighter coloring indicates models which only have a single pathology assessment, while darker coloring indicates models which have Aβ, tau, and neurodegeneration biomarkers. Gold stars indicate a significant improvement in accuracy relative to the topmost model.

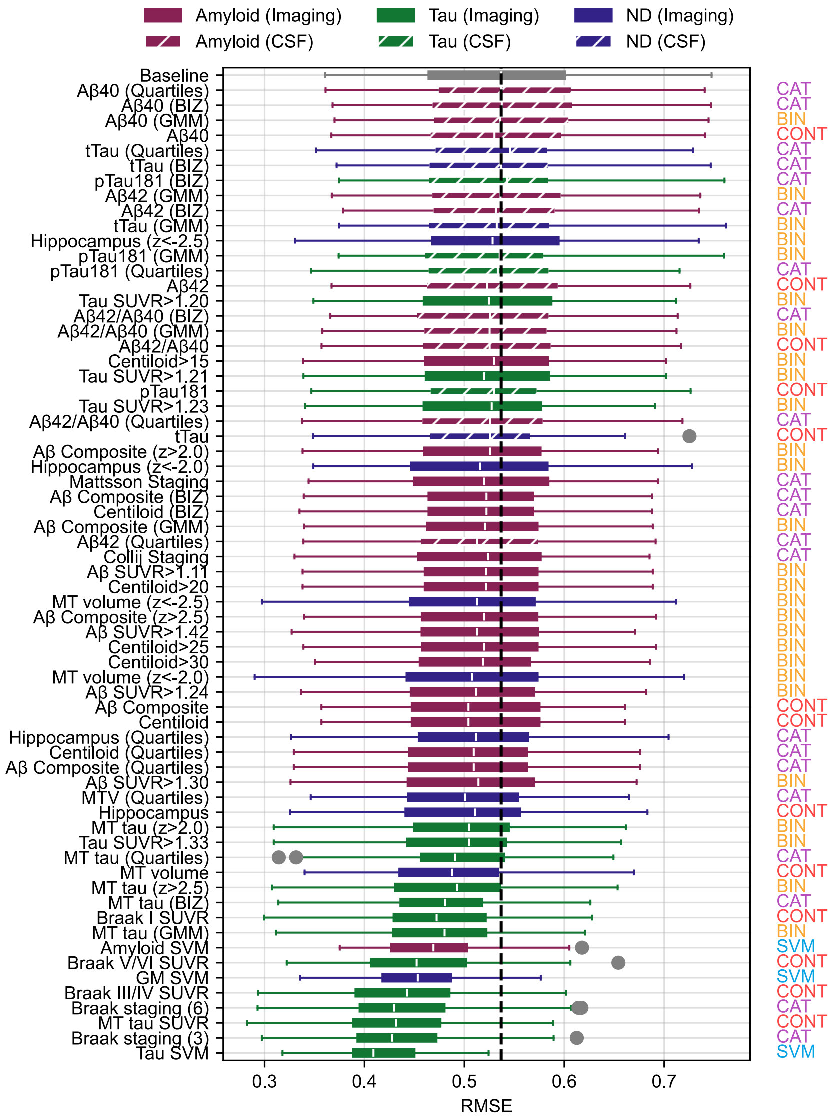


**eFigure 7**: Alternate version of main text Figure 1 including CSF predictors alongside imaging-based ones.
